# Correlation between Mid-Upper Arm Circumference and Body Mass Index in the assessment of adults’ nutritional status in Malawi

**DOI:** 10.1101/2024.07.09.24310002

**Authors:** Thokozani Mzumara, Adriano Focus Lubanga, Joseph Afonne, George Munthali, Byenala Kaonga, Gracian Harawa, Akim Nelson Bwanali

## Abstract

**Background:** Body Mass Index (BMI) is a widely used and accepted indicator of nutritional status in adults. Mid-Upper-Arm-Circumference (MUAC) is another anthropometric measure used primarily among children. While BMI remains the best indicator of nutritional status, it can sometimes be impractical because of logistical requirements for weight and height measurement, especially for large population-based studies and bed-ridden patients. Therefore, we analyzed anthropometric data collected from the Malawi Epidemiology and Intervention Research Unit (MEIRU) Non-Communicable Disease (NCD) survey to determine the correlation between BMI and MUAC in the assessment of adult nutritional status in Malawi.

**Methods:** A secondary data analysis utilizing descriptive and correlational statistical research methods was used to determine the relationship between BMI and MUAC in the assessment of adults’ nutritional status in Malawi. The data was analyzed using SPSS version 27. The independent t test and Chi-square were employed. Furthermore, the study included the Pearson correlation test to assess the relationship between variables. A p-value of <0.05 was considered statistically significant.

**Results:** The study assessed 30,575 participants, of whom the majority (61.8%) were females. The mean MUAC was 27.2 (SD = 3.300), and the mean BMI was 23.5 (SD = 4.55). The study found a strong positive statistically significant correlation between MUAC and BMI among Malawians (r = 0.836, CI = (0.832-0.839) such that for each additional centimeter increase in MUAC, BMI is expected to increase by approximately 1.153 units (BMI = -7.797 + 1.153 (MUAC)). There was a significant positive correlation between BMI and MUAC in both males and females and in rural and urban residents (P<0.01). The ROC curve was excellent for BMI in the overweight range (AUC = 0.87), and the findings were superior in the obese range (AUC = 0.956).

**Conclusion:** The correlation between MUAC and BMI is positive regardless of sex or rural/urban residence. Therefore, the MUAC can be used as a clinical test to predict BMI.

## Introduction

Worldwide, around 890 million adults are living with obesity, while 390 million are underweight. In sub-Saharan Africa (SSA), the overall prevalence of overweight/obesity among adults in the region was estimated to be 42% [1]. Recently, Malawi has experienced the rise of obesity and overweight [2], and literature suggests that about 21% of Malawian adult women are overweight or obese [3]. A study found that overweight and/or obesity are the most common public health issues in Malawi, impacting at least one in every five people [4]. The condition affects more women than men, and it is more prevalent in cities than in rural areas [4]. As a consequence, an increase in weight is strongly linked to a rise in waist-hip ratio, a proven risk factor for NCDs such as diabetes [5].

Malnutrition refers to deficiencies, excesses, or imbalances in a person’s intake of energy and/or nutrients [6]. Generally, it covers 2 broad groups of conditions. One is ‘undernutrition’—which includes stunting (low height for age), wasting (low weight for height), underweight (low weight for age), and micronutrient deficiencies or insufficiencies (a lack of important vitamins and minerals) [7]. The other is overweight, obesity, and diet-related non-communicable diseases (NCDs) such as heart disease, stroke, diabetes, and cancer [8]. BMI is a widely used indicator of nutritional status in adults, while the MUAC is another anthropometric measure used primarily among children. MUAC is a viable alternative to BMI for screening adult underweight. The MUAC is a measurement of the upper arm circumference at the halfway point between the elbow tip (olecranon process) and the shoulder blade tip (acromion process) [9]. While MUAC measurements are often a reflection of both muscle and subcutaneous fat, in undernourished people with lower quantities of subcutaneous fat, MUAC values can indicate a chronic energy shortage. MUAC readings are linear and may be obtained with a simple tape measure. With suitable MUAC cut-offs, anybody with minimal training can perform the examination using a simple paper strip with color-coded cut-off values [9].

BMI and MUAC stand as pivotal anthropometric indicators utilized in assessing nutritional status. In general, BMI is widely recognized as a standard measure of adiposity and nutritional status, MUAC offers a convenient and reliable alternative, particularly in resource-constrained settings [10]. Nevertheless, the measurement of the MUAC offers a simple and sensitive method for early diagnosis of at-risk groups of both overweight and underweight [11]. According to Brito and colleagues [12], MUAC can represent a useful measurement of malnutrition, especially in the absence of weight and height measurements, which constitute BMI [13].

Despite the prominence of BMI and MUAC in nutritional assessment, limited comprehensive studies have explored the intricate relationship between these two metrics among populations in developing countries such as Malawi. Understanding this relationship is essential for more accurate and nuanced assessments of nutritional status, enabling targeted interventions to combat malnutrition effectively. By conducting a thorough analysis, encompassing diverse age groups, genders, and geographical locations, this study seeks to shed light on MUAC as a proxy measurement for BMI in the Malawian context. There is an urgent need to establish population-based information on MUAC and BMI to ensure effective screening in low-resource areas such as outreach or field clinical investigations where time and resources, including literacy and numeracy skills, are involved to a certain extent [14]. This approach necessitates considerable training and the use of calibrated medical weighing scales and stadiometers [15].

By unraveling these mysteries, we hope to provide valuable insights into the nutritional landscape of Malawi and inform evidence-based policies and interventions aimed at improving nutritional outcomes and overall population health. In addition, through statistical analysis, and interpretation, this study endeavors to contribute to the growing body of literature on nutritional assessment methodologies while offering practical implications for public health practitioners, policymakers, and researchers working in the field of nutrition and health in Malawi and beyond. Therefore, this study aimed to comprehensively examine the relationship between BMI and MUAC among Malawians across sex and rural-urban residence strata.

## Materials and Methods

### Study design and setting

The cross-sectional quantitative study adopted a secondary research technique. Data for the study was retrieved from a survey conducted by MEIRU between May 2013 and April 2017. The original data was collected from two sites in Malawi, namely Karonga in the Northern Region of Malawi and Area 25 in Lilongwe, the Central Region of Malawi. Lilongwe is the capital city and the largest city in the country [16]. Area 25 represents an economically mixed urban economy with a population of about 66,000 people, and Karonga is a lakeshore area with a subsistence economy and a rural area [17]. The data utilized questionnaires administered through an electronic platform. Some data was obtained from laboratory results.

### Inclusion and exclusion criteria

The study included all adults aged 18 and above who resided in the house and excluded all adults who identified as visitors.

### Sampling procedure

The study population and sampling frame are described in a previous study [18]. The study population was made up of people who took part in the Malawi Epidemiology and Intervention Research Unit Noncommunicable Disease Survey, which was conducted between 2013 and 2017. The study focused on residents from various regions of Malawi, to determine the prevalence and risk factors for noncommunicable diseases. The sampling frame was developed to ensure a representative sample of the population by considering a varied range of demographic and geographical criteria.

### Data Management

BMI was recoded into categorical variables, classifying the participants into 4 categories based on BMI measurements. The subject’s BMI determines their classification as underweight (BMI < 18.5 kg/m2), within the normal range (BMI = 18.5 to 24.9 kg/m2), overweight (BMI > 24.9 to 29.9 kg/m2), or obese (BMI ≥ 30 kg/m2) [12].

### Data Analysis

The data were analyzed using IBM Statistical Package for Social Sciences (SPSS) Statistics version 27. Descriptive statistics were generated for demographic variables, all measurements (weight, height, and MUAC), and BMI. A curve estimation was done to assess the linearity of BMI and MUAC. Subsequently, a correlation analysis between BMI and MUAC was performed. Pearson correlation was used to assess the correlation between variables. Furthermore, linear regression analysis was conducted to predict BMI values from MUAC. The ROC curve was used to set cutoff values, with the optimal area being less than 0.1-0.9. Accordingly, AUC ranges from greater than 0.9 depict excellent model discrimination [19]. Results with a p-value of < 0.05 were considered statistically significant.

### Ethical consideration

Due to the nature of the study, we did not require an ethical review since the data used is freely available online upon request. However, the initial study was ethically approved (Ref number: IRB No. 13/2012) and adhered to the Declaration of Helsinki.

## Results

The study assessed 30575 participants, of whom the majority (61.8%) were females (Table 1). The majority of participants were from urban areas (54.5%). Most of the participants were aged 15– 34 years (18263) (59.7%) and the least number of participants were aged 55–64 (3588) (11.7%) (Figure 1)

**Figure 1.**
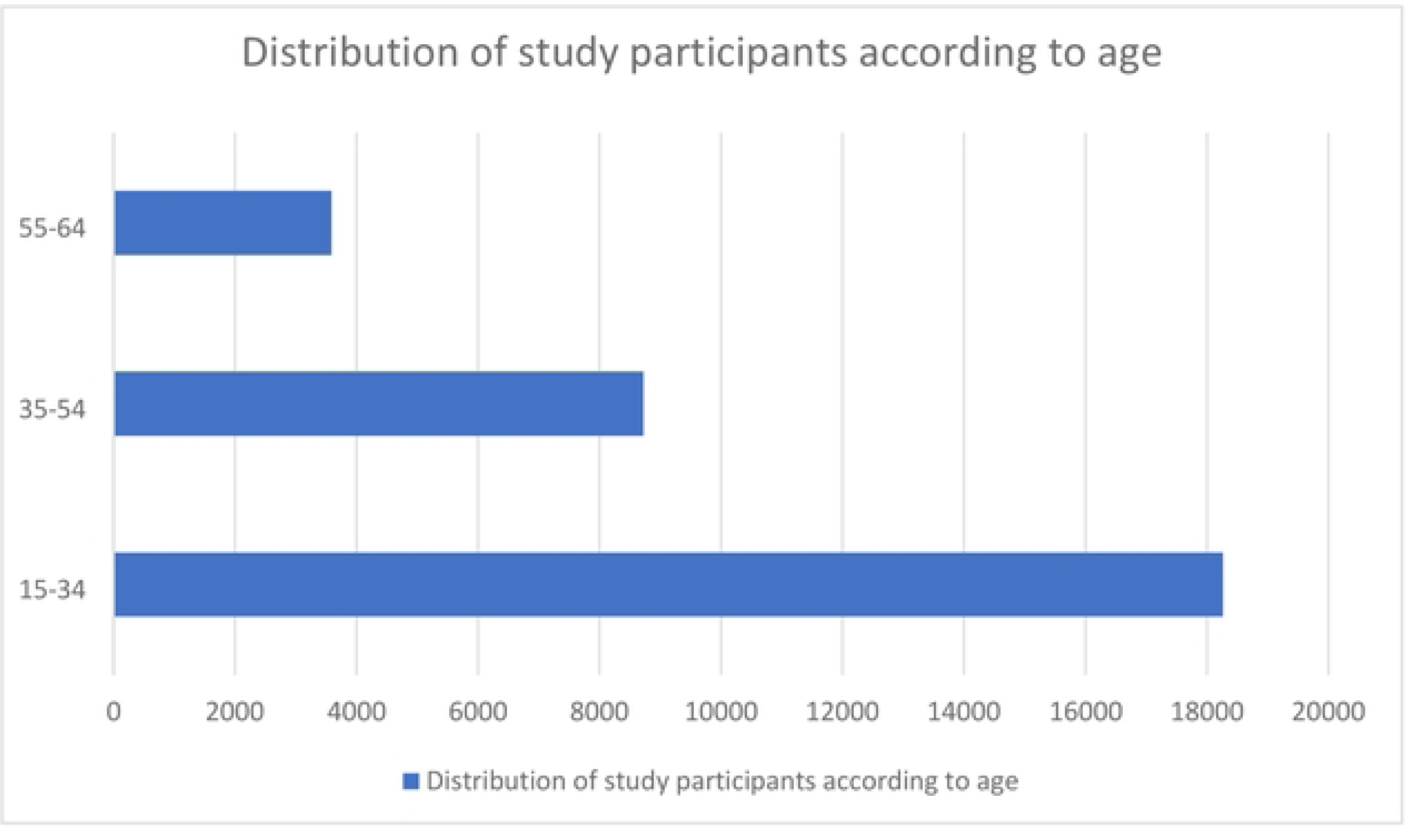
Distribution of study participants according to age

**Table 1.** Demographic characteristics of study participants.

| **Gender** | **Number** | **Percentage (%)** |
| --- | --- | --- |
| Male | 11668 | 38.2 |
| Female | 18907 | 61.8 |
| **Location** |  |  |
| Urban | 16672 | 54.5 |
| Rural | 13903 | 45.4 |

Based on BMI, most participants were within the normal range, 20674 (67.6%), and the least number of participants were underweight, 1757 (5.7%). Based on gender, more males were underweight compared to females. And those in rural areas had a higher prevalence of underweight than those in urban areas. However, considering overweight and obesity, the prevalence was higher among females and those in urban areas (Table 2).

**Table 2.** Interpretation of Weight Based on Body Mass Index according to gender and rurality.

|  | **TOTAL** | **Male** | **Female** | **Rural** | **Urban** |
| --- | --- | --- | --- | --- | --- |
| **Underweight** | 1757 (5.7) | 897(7.6) | 860 (4.5) | 1009 (7.3) | 748 (4.5) |
| **Normal** | 20674 (67.6) | 9176 (78.6) | 11498 (60.8) | 10354 (74.5) | 10320 (61.9) |
| **Overweight** | 5442 (17.8) | 1312 (11.2) | 4130 (21.8) | 1909 (13.7) | 3533 (21.2) |
| **Obese** | 2702 (8.8) | 283 (2.4) | 2419 (12.8) | 631(4.5) | 2071(12.4) |

### Distribution of MUAC and BMI according to gender

The mean BMI score was 23.5 kg/m² (SD = 4.55). According to gender, males 21.99 kg/m² (SD = 3.20) participants had a lower BMI compared to females 24.54 kg/m² (SD = 5.001). An independent t-test found that the difference in mean BMI-based rurality was statistically significant (t(29448) = -48.765, p <0.001).

The mean MUAC was 27.2 cm (SD = 3.300). According to gender, males had a lower MUAC of 26.45 cm (SD =2.682) than females at 27.60 cm (SD = 3.55). An independent t-test found that the difference in mean MUAC-based rurality was statistically significant (t(30573) = -30.128, p <0.001).

### Distribution of MUAC and BMI according to location

According to location, the average BMI was higher in Lilongwe at 24.33 kg/m² (SD = 4.96) than in Karonga at 22.57 kg/m² (SD = 3.788). An independent t-test found that the difference in mean BMI-based rurality was statistically significant (t(29448) = 33.677, p <0.001). Likewise, MUAC was higher in Lilongwe at 27.69 cm (SD = 3.39) than in Karonga at 26.52 cm (SD = 3.07). An independent t-test found that the difference in mean MUAC-based rurality was statistically significant (t(30573) = 31.33, p <0.001).

### Relationship between MUAC and BMI

The Pearson correlation test indicated a statistically significant correlation between MUAC and BMI (r = 0.836, CI = 0.832–0.839) (figure 2) A simple linear regression was used to estimate body BMI from MUAC. A significant regression equation was found (F(1, n-2) = 68334, p < 0.001), with a R² of 0.699. This means that MUAC can explain approximately 69.9% of the variation in BMI. When MUAC is measured in centimeters, the projected BMI of the individuals is -7.797 + 1.153 (MUAC). BMI increased by 1.153 per centimeter of MUAC. MUAC was a strong predictor of BMI (r = 0.836, p < 0.001).

**Figure 2.**
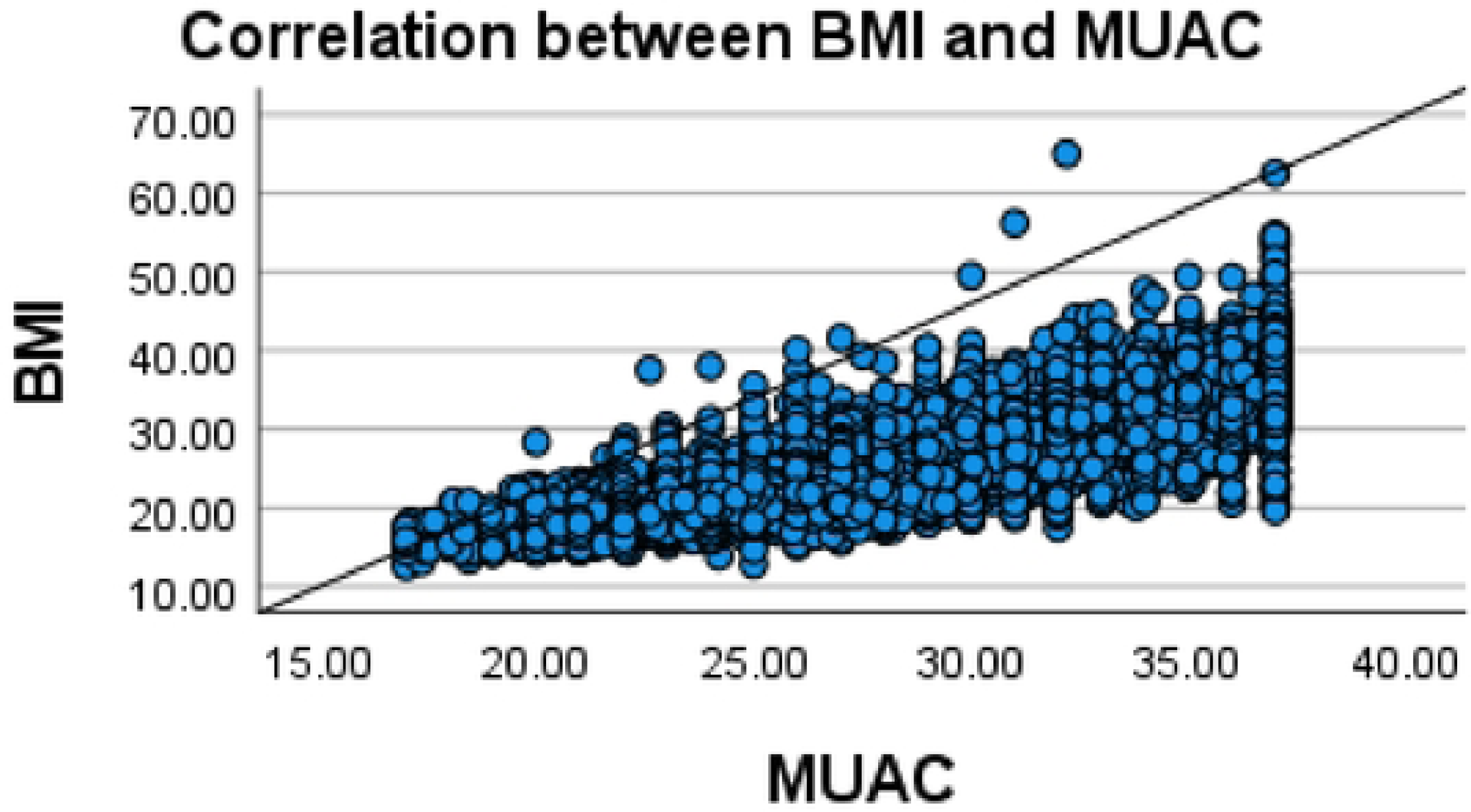
Correlation between BMI and MUAC

### Relationship between MUAC and BMI according to sex

The Pearson correlation coefficient test showed that there was a positive correlation between BMI and MUAC among male participants, and the correlation was statistically significant (r= 0.759, p<0.001 (CI = 0.751, 0.766).

**Figure 3.**
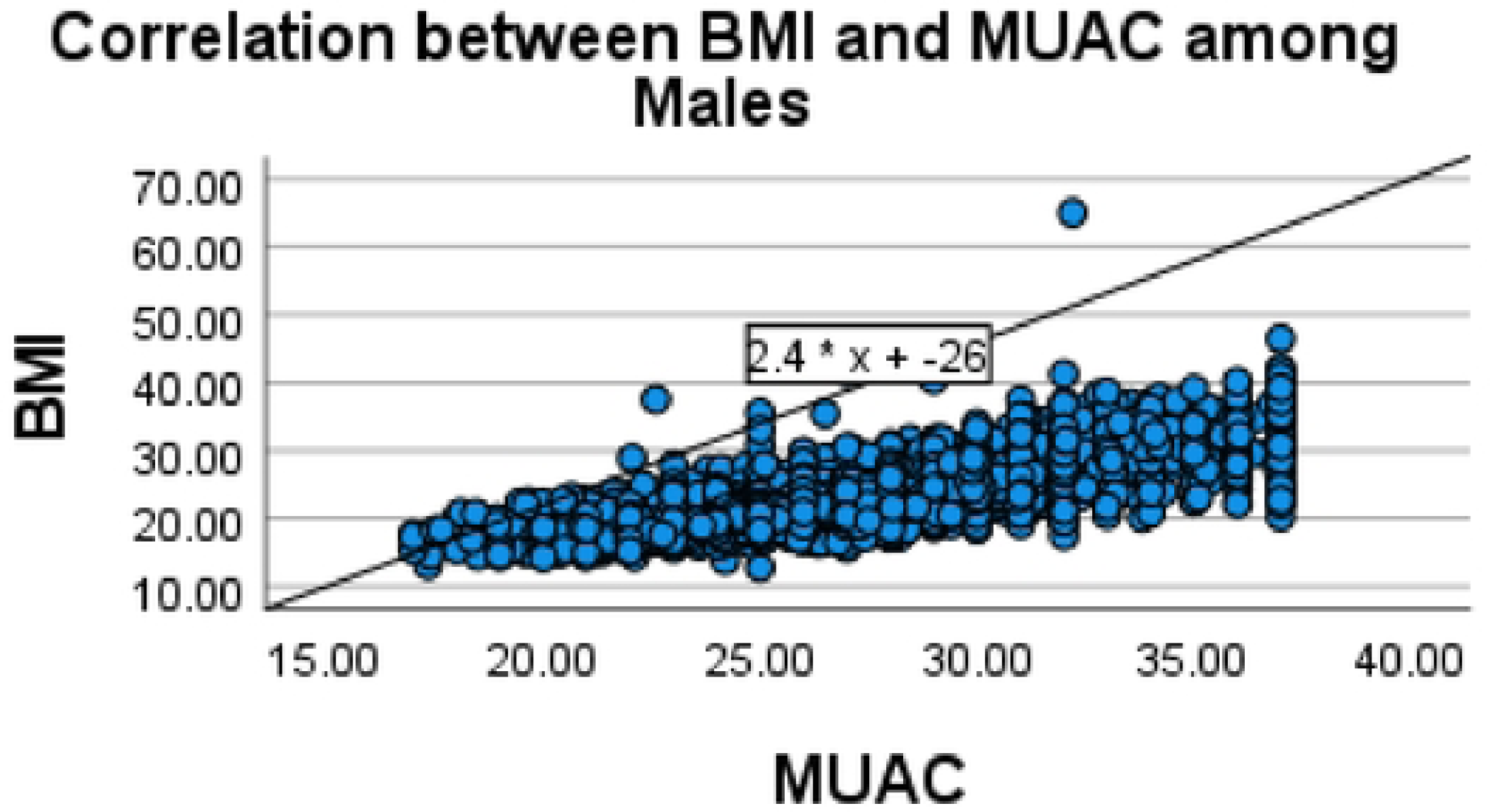
Correlation between BMI and MUAC among males

As shown in Figure 4 below, the Pearson correlation test showed that there is a statistically significant positive correlation between BMI and MUAC among females (r = 0.857, p<0.001, CI = 0.853, 0.861).

**Figure 4.**
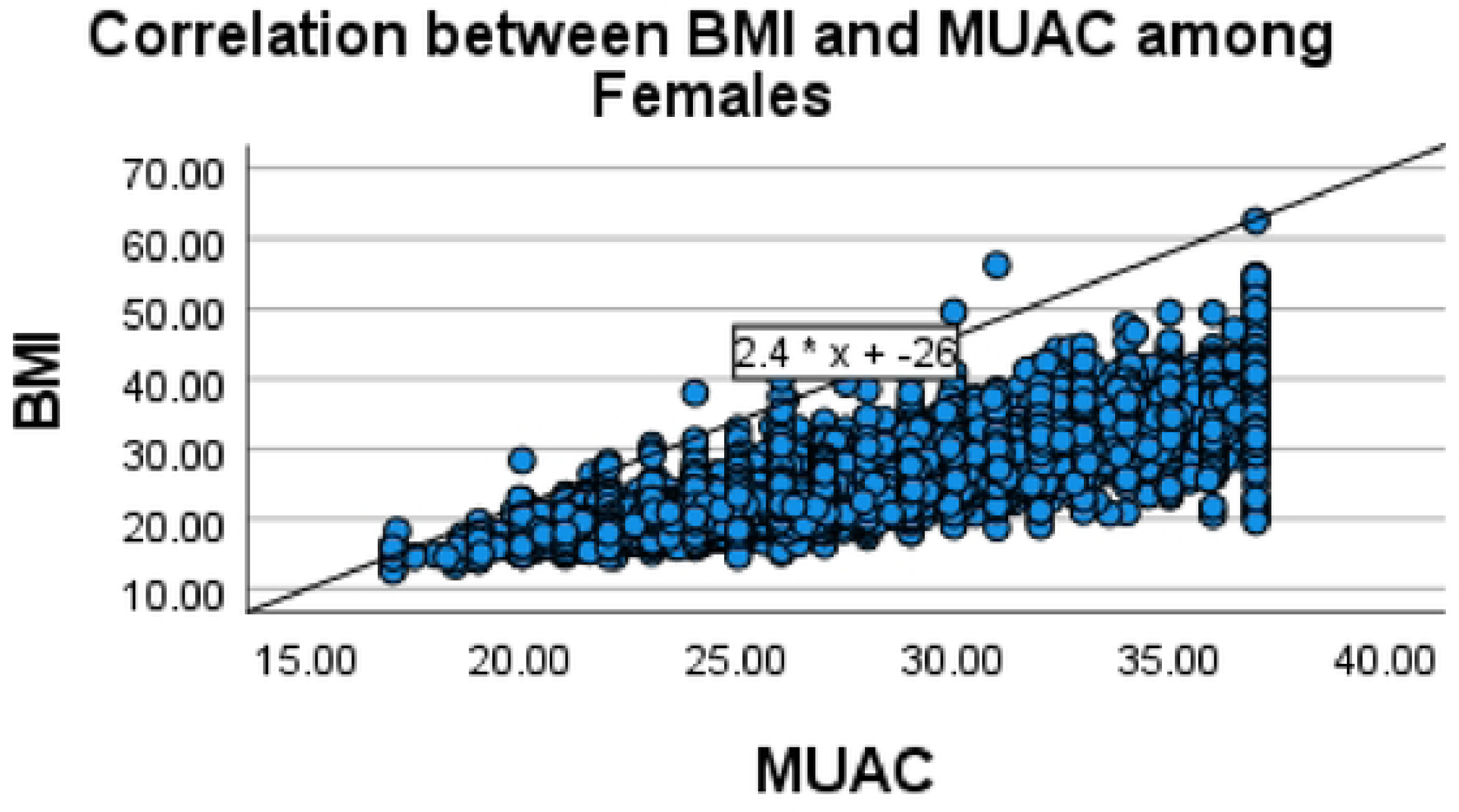
Correlation between BMI and MUAC among female

### Relationship between MUAC and BMI according to rurality

The Pearson chi-square showed a positive correlation that was statistically significant (r = 0.820, p<0.001), as shown in Figure 5.

**Figure 5.**
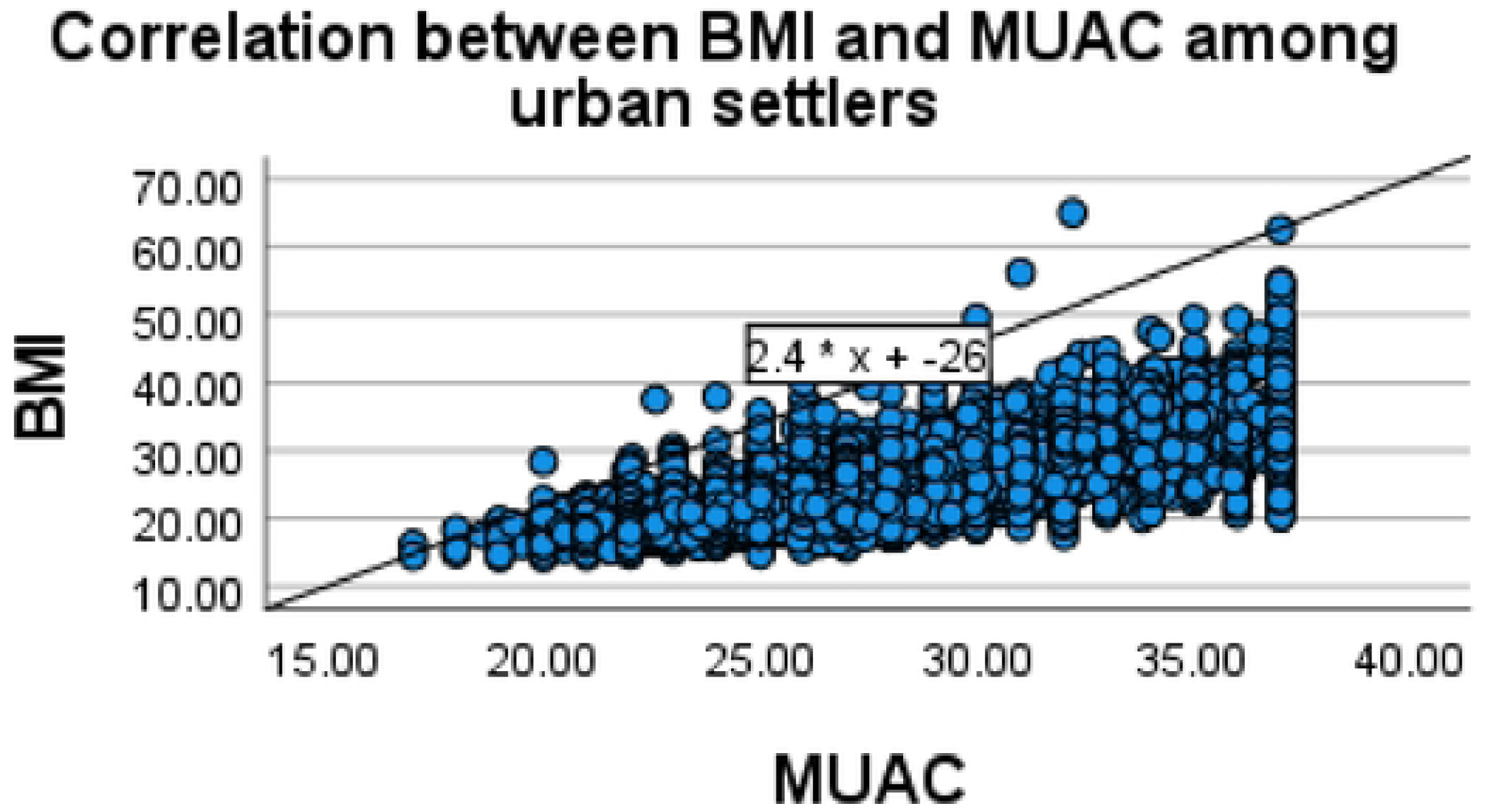
Correlation between BMI and MUAC among urban settlers

According to Figure 6 below, the Pearson correlation coefficient test showed that the relationship between MUAC and BMI among rural settlers was positive and statistically significant (r = 0.856, p<0.001).

**Figure 6.**
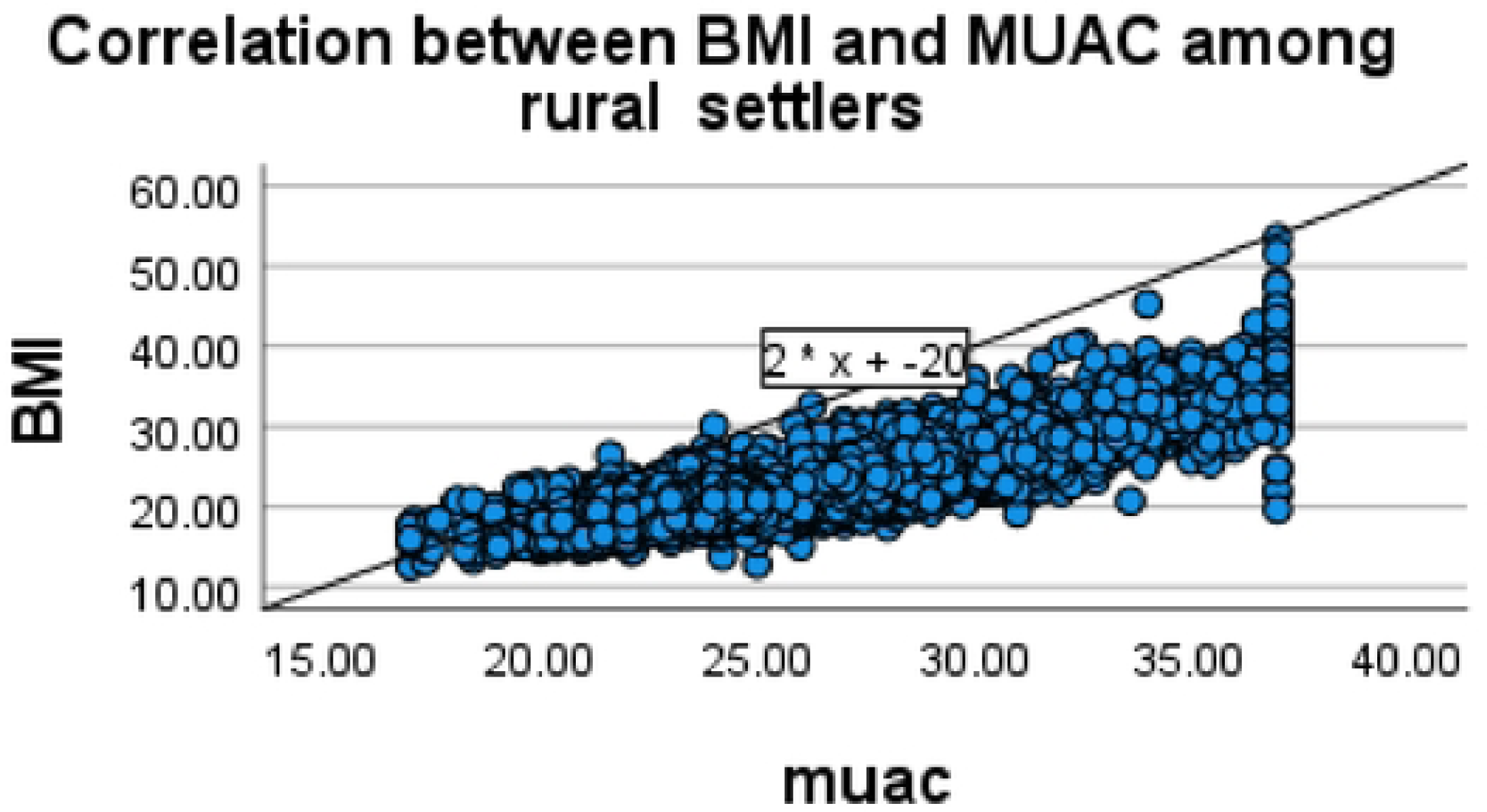
Correlation between BMI and MUAC among rural settlers

### Determining MUAC cut-off points

The area under the ROC curve for underweight (Figure 7) and normal weight (Figure 8) was 0.076 and 0.232, respectively. Nevertheless, the ROC curve was excellent for BMI in the overweight range (AUC = 0.87), and the findings were superior in the obese range (AUC = 0.956).

**Figure 7.**
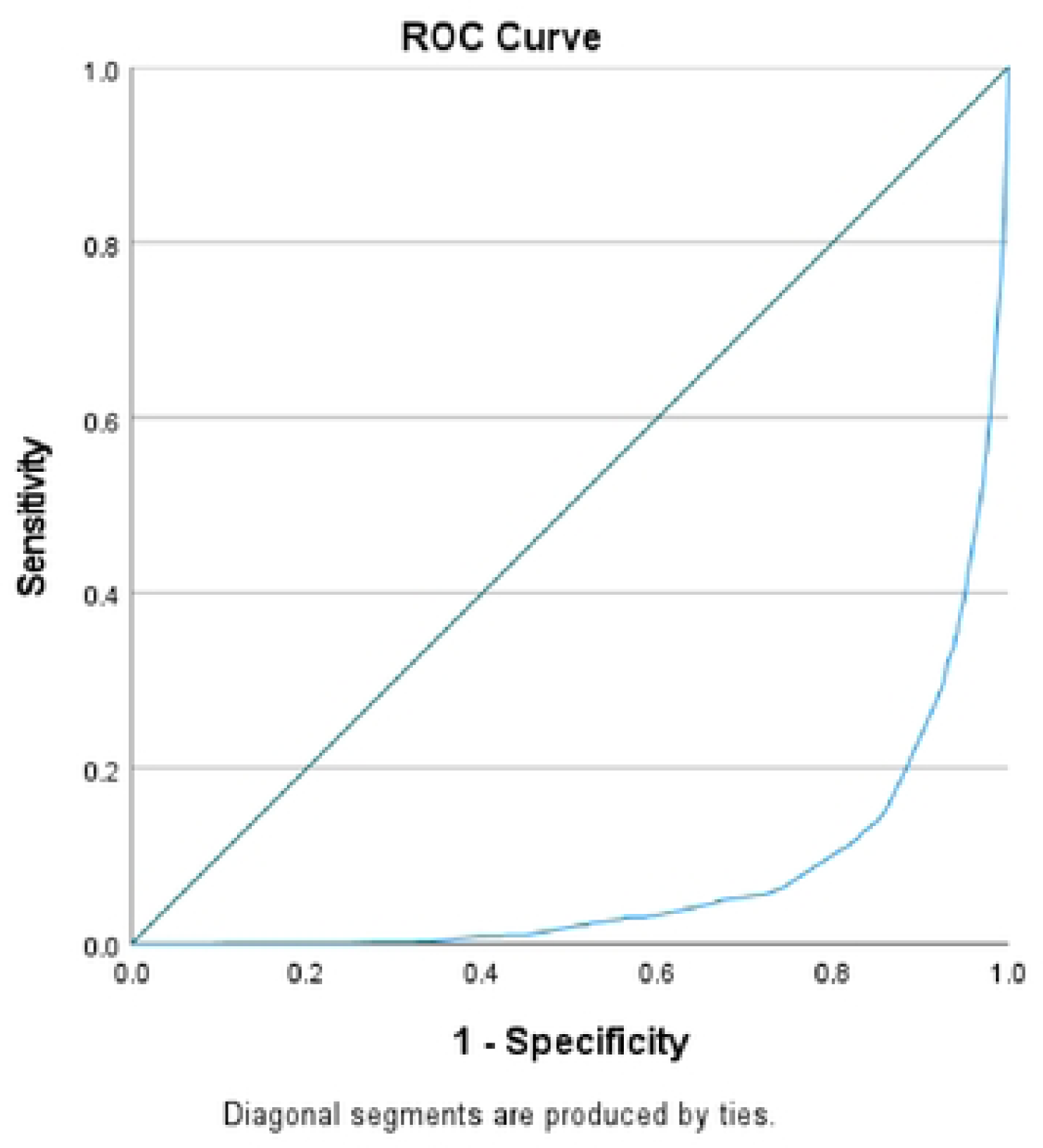
ROC Curve for Underweight

**Figure 8.**
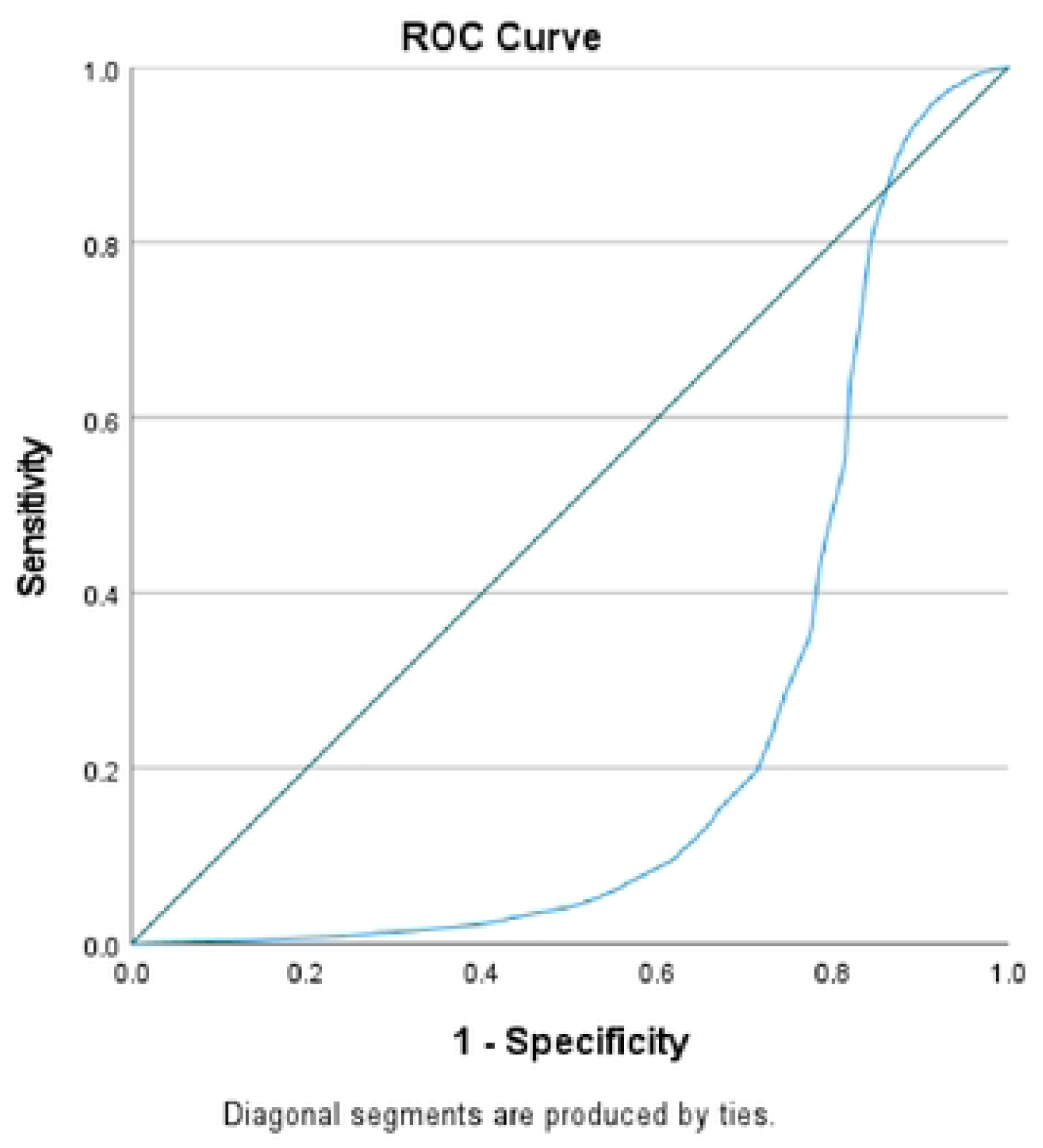
ROC of Normal

**Figure 9.**
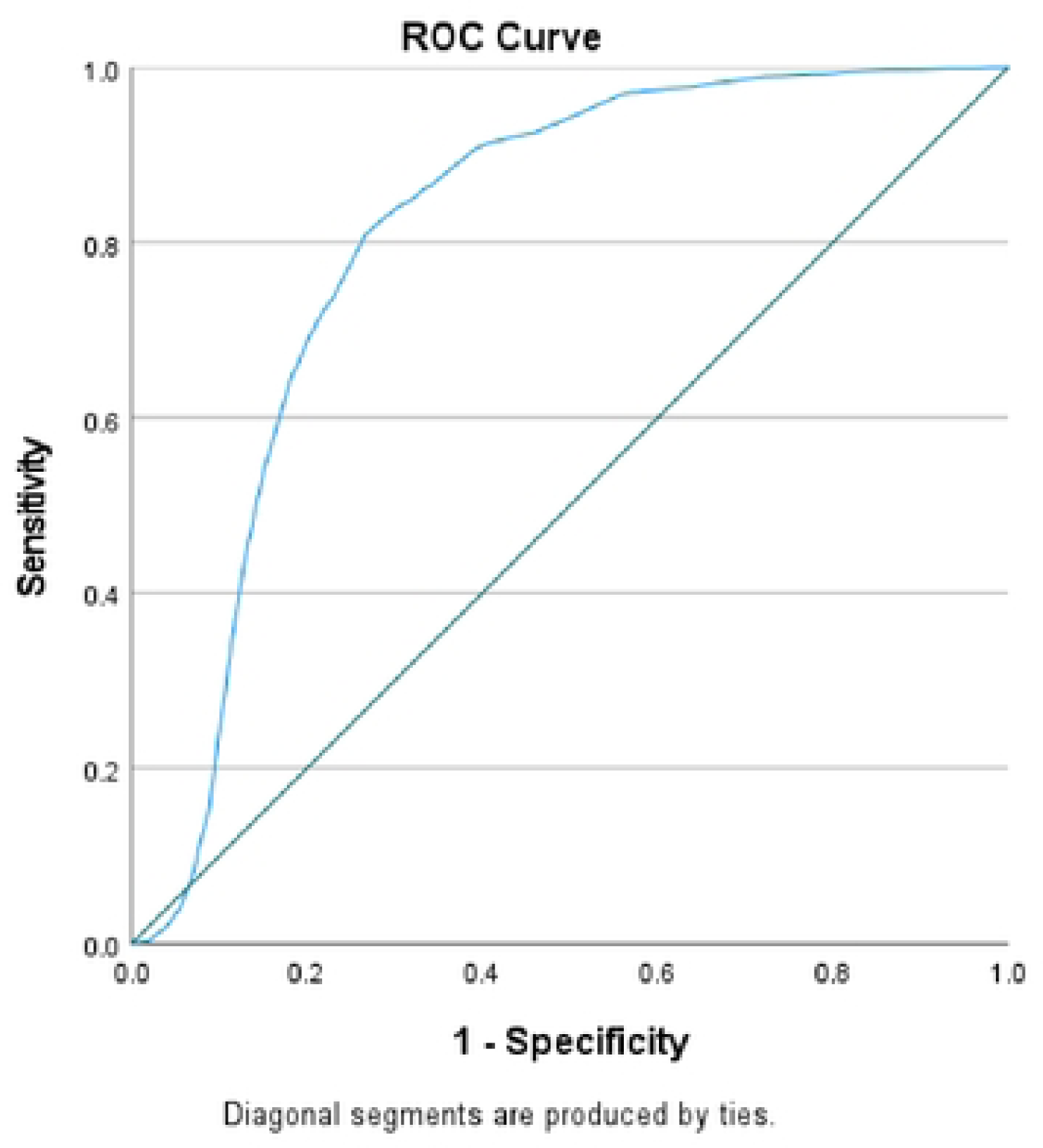
ROC OF OVERWEIGHT

**Figure 10.**
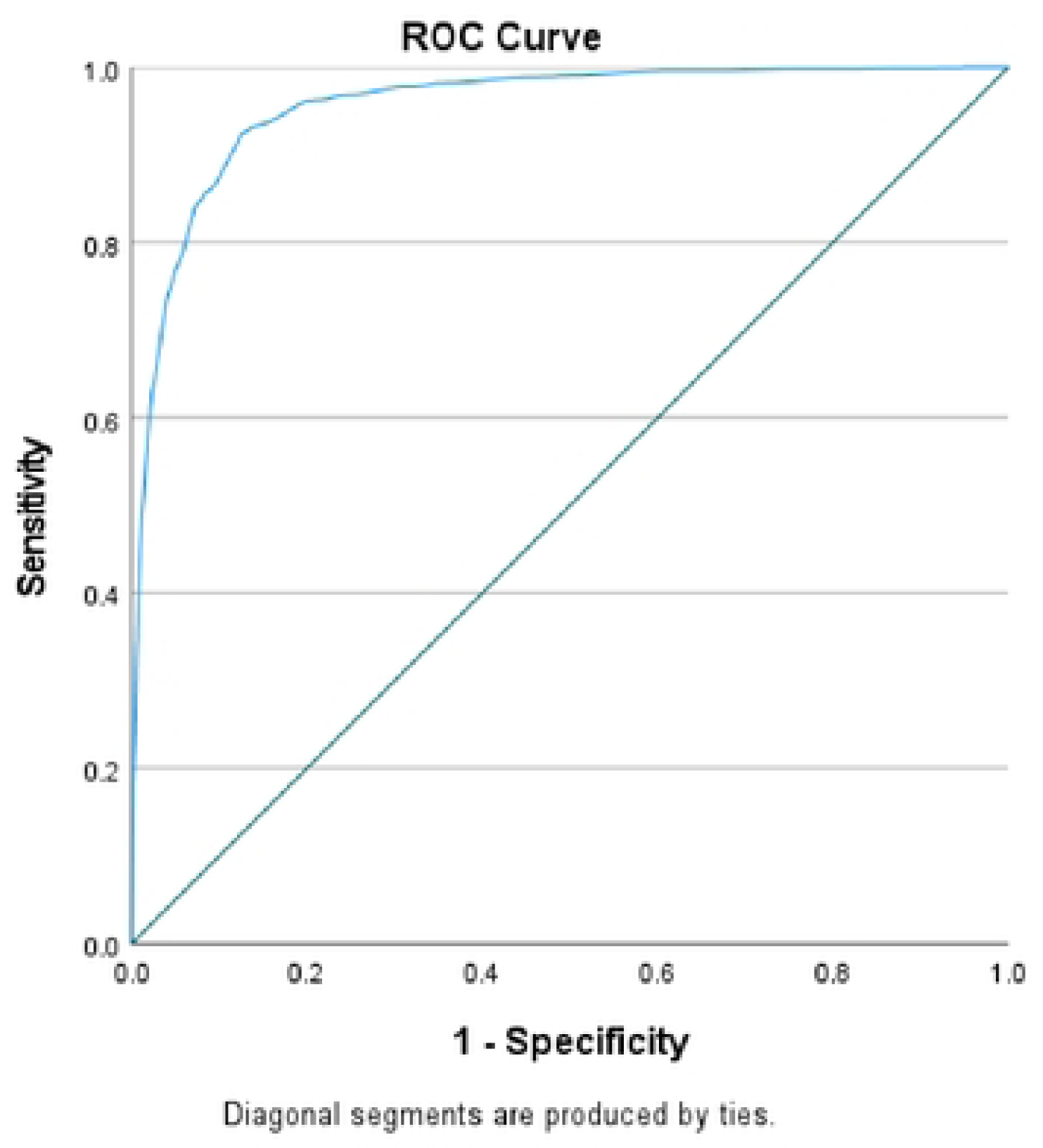
ROC Curve for Obesity

## Discussion

The study aimed to assess the relationship between MUAC and BMI among the Malawian population. In the current paper, MUAC and BMI were strongly correlated, and the correlation was statistically significant, confirming that MUAC can be used as an alternative to BMI measurement among this population group.

The study reports a positive correlation between BMI and MUAC similar to a study conducted among university students in Pakistan [20], a study conducted among students in South Africa [21], and other studies conducted in Sudan [22] and elsewhere [23, 24]. Furthermore, this study finding is similar to Brito and colleagues [12], who found a statistically positive correlation between MUAC and BMI among inpatients. The study findings add to the existing literature that suggests a positive relationship between MUAC and BMI. In particular, this study is the first one conducted among this population group. The findings of this study highlight the fact that MUAC can be used to detect underweight, overweight, and obesity among the study population. Developing countries such as Malawi are currently facing the double burden of NCD, with a rise in overweight and obesity [25]. This study’s findings, if incorporated into practice, provide a simple and efficient method of screening suitable for outreach programs, including bedridden patients. Furthermore, this finding is critical in the context of task-shifting policies that allow less specialized cadres, such as community health workers and hospital attendants, in hard-to-reach areas to conduct malnutrition status checks. Task-shifting approaches have been proven to be an efficient health management technique in light of the shortage of healthcare workers [26].

The study found that for each additional centimeter increase in MUAC, BMI is expected to increase by approximately 1.153 units. This finding can be particularly useful in clinical settings where MUAC can be used as a quick and reliable measure to estimate BMI and assess nutritional status. In Africa, two-thirds of patients are at risk of developing malnutrition issues [27]. Similarly, Malawi reports high rates of malnutrition, calling for comprehensive screening practices [27].

Determining the MUAC cut-off point from the ROC curve was not significant in people with underweight or normal weight, but it was significant in the overweight and obese range, similar to a previous study [28]. In contrast, others [29] found that MUAC was a good predictor for underweight. The difference in results can be attributed to variations in geographical areas and study populations. The study recommends implementing MUAC cut-off values based on regional data, conducting validation studies, and integrating them into national health protocols. These measures aim to improve nutritional assessments, reduce health disparities, and strengthen health systems.

Although both genders reported positive correlations, females demonstrated a stronger correlation compared to men, similar to previous authors [11]. This study’s finding reflects gender differences in the parameters due to genetic and biological factors such as the effect of sex hormones [30]. Although the correlation between MUAC and BMI was positive and significant in both rural and urban areas, there was a slight difference between the two sites, which are urban and rural. This agrees with a previous paper [11], that reported stronger correlations between MUAC and BMI in Tanzania as compared to Mozambique. The study findings postulate that the relationship between MUAC and BMI may vary based on geographical location. Geographical disparities related to noncommunicable diseases in rural and urban areas exist due to changes in demography and structure [30].

The average MUAC and BMI in this study are in tandem with previous studies [12]. Nevertheless, according to gender, females had a higher BMI and a higher MUAC. This is not surprising considering that obesity and overweight in developing countries are major problems facing adult women, unlike men [31]. In Malawi, the prevalence of obesity among women has doubled, from 10% in 1992 to 21% in 2016 [2]. The findings of this study can be attributed to the fact that among Malawian women, gaining weight is strongly linked with doing well in life; hence, whenever resources allow, women strive to gain weight to attain a symbol of wealth and improve their personal dignity [32]. Although the study did not assess the factors associated with being overweight, the literature indicates that women, especially those in marriage, view being overweight as a sign of opulence and wealth, as large body size is viewed in light of successful marriage among women [30]. Besides behavioral factors such as physical inactivity, genetic and biological factors also play a major role for women. The findings of this study highlight the need to streamline strategies to curb overweight by considering wealth, gender, and marital status as risk factors for overweight.

In the study, participants in urban areas had a higher BMI than those residing in rural areas, on par with a recent study conducted in the country, which found that poverty is linked with the odds of being underweight [2]. Malawi is a country currently going through rapid urbanization [33]. Consequently, urbanization is strongly linked to a sedentary lifestyle and easy access to unhealthy foods (high in calories and low in dietary fiber), which lead to overweight and obesity [30]. Furthermore, the nutritional status of a country is strongly linked to levels of economic development, gross domestic product, and urbanization [34]. Needless to say, overweight and obesity in Malawi can be explained by both individual and community factors [32]. Furthermore, one of the major contributing factors to malnutrition in Malawi is the lack of awareness regarding the dangers of gaining weight through various media outlets, including social media platforms [2]. This finding can be attributed to the increased economic activities and income-generating activities associated with city life, where people may manage to use disposable income on extra food, leading to overweight. For instance, Ndambo and colleagues [32] found that Malawians who have a normal weight were those who admitted to eating less than they would like to owing to financial constraints. Another study found that the risk of overweight and obesity increases with increasing weight among the Malawian population [35]. In addition, underweight could be attributed to increased physical activities in the rural areas associated with farming activities, unlike the urban sedentary lifestyles fueled by office work and motorized transportation in the city. The findings of this study suggest efforts to combat obesity should focus on the need for regular exercise, healthy eating behaviors, and public awareness campaigns as viable management options for non-communicable diseases in urban areas.

## Limitations

The study is not without limitations. Firstly, the study does not represent a nationally representative survey and hence cannot be generalized to a wider Malawian context considering that only segments of two districts were covered in the analysis. Secondly, the study does not represent all age groups, especially children. There was also a lack of data regarding morbidities such as HIV/AIDS and tuberculosis. In addition, the study did not classify the four categories of nutrition status based on MUAC, as it is beyond the scope of the study. Nevertheless, this study is the first to be conducted on the topic among the population group.

## Conclusion

In conclusion, this study contributes to the literature on adult nutrition in a resource-limited setting. Given that the study found that the correlation between MUAC and BMI is strong and positive regardless of gender and rural/urban residence, the study recommends that MUAC can be used as a simple and low-resource nutrition screening tool among this population. Hence, the study findings contribute to the fight against NCDs in the country by promoting early diagnosis and management of NCDs in Malawi. It recommends training and education on MUAC among low-level cadres to increase coverage of NCD screening in rural and urban areas. More studies are required to further understand this phenomenon, targeting various high-risk population groups such as children, pregnant women, and people living with HIV/AIDS. In addition, a nationwide study is needed to elucidate the concept beyond two areas.

## Declarations

## Acknowledgments

None

## Data availability

The data utilized for this study is freely accessible online and can be accessed via the following link https://doi.org/10.17037/DATA.00000961

## Informed consent

Not applicable

## Ethical approval

Due to the nature of our research work, the ethical approval was waived.

